# Delayed Stroke Treatment during COVID-19 Pandemic in China

**DOI:** 10.1101/2020.11.17.20228122

**Authors:** Shiyuan Gu, Zhengze Dai, Huachao Shen, Yongjie Bai, Xiaohao Zhang, Xinfeng Liu, Gelin Xu

## Abstract

**Background:** Social distance, quarantine, pathogen testing and other preventive strategies implemented during COVID-19 pandemic may negatively influence the management of acute stroke.

**Objective:** The current study aimed to evaluate the impacts of COVID-19 pandemic on treatment delay of acute stroke in China.

**Methods:** This study included patients with acute stroke admitted in two hospitals in Jiangsu, China. Patients admitted before and after the COVID-19 epidemic outbreak (January 31, 2020, as officially announced by Chinese government) were compared for pre- (measured as onset-to-door time) and post-hospital delay (measured as door-to-needle time). The influence factors for delayed treatment (indicated as onset-to-needle time >4.5 hours) were analyzed with multivariate logistic regression analysis.

**Results:** Onset-to-door time increased from 202 min (IQR 65-492) before to 317 min (IQR 75-790) after the COVID-19 pandemic (*P=*0.001). Door-to-needle time increased from 50min (IQR 40-75) before to 65 min (IQR 48-84) after the COVID-19 pandemic (*P=*0.048). The proportion of patients with intravenous thrombolysis in those with acute ischemic stroke was decreased significantly after the pandemic (15.4% vs 20.1%; *P*=0.030). Multivariate logistic regression analysis indicated that patients after COVID-19 pandemic, lower educational level, rural residency, mild symptoms and transported by other means than ambulance were associated with delayed treatment.

**Conclusions:** COVID-19 pandemic has remarkable impacts on the management of acute ischemic stroke. Both pre- and post-hospital delays were prolonged significantly, and proportion of patient arrived within the 4.5-hour time window for intravenous thrombolysis treatment was decreased. Given that anti-COVID-19 measures are becoming medical routines, efforts are warranted to shorten the delay so that the outcomes of stroke could be improved.

## Introduction

Recently developed treatments, such as intravenous thrombolysis and mechanical thrombectomy, can significantly improve the outcomes of acute ischemic stroke. But the effects of these treatments were highly time-dependent, which emphasize the importance of rapid pre- and post-hospital managements. For selected patients with onset-to-needle time (ONT) shorter than 4.5 hours, intravenous thrombolysis could be applied. But those with ONT shorter than 3 hours had a higher likelihood of 90-day favorable outcome [1]. For selected patients with onset-to-puncture time (OPT) shorter than 6 hours, mechanical thrombectomy could be applied. Although patients with OPT between 6 and 24 hours still could be screened for mechanical thrombectomy, the effects attenuate rapidly with time delay. Current guidelines recommended that extra imaging examinations should be performed for selecting patients with OPT between 6 and 24 hours for mechanical thrombectomy [2,3]. Therefore, when applying intravenous thrombolysis and mechanical thrombectomy in acute ischemic stroke patients, the shorter the treatment delays, the better the functional outcomes.

Since the outbreak of COVID-19 pandemic, China has implemented several nation-wide strategies for preventing and containing the spread of the disease [4]. Social distance, quarantine, pathogen testing and other strategies were taken from January 31, 2020, as officially announced by Chinese government. These measures influenced not only the regular medical procedures, but also the health-seeking behaviors. All these changes may have influenced the management of stroke, but the impacts are largely undetermined [5]. This study aimed to explore the impact and extent of COVID-19 pandemic on treatment delay of acute stroke in China. Additionally, we probed potential factors responsible for the treatment delay.

## Methods

### Study design and patient population

This study is a part of an on-going program for analyzing pre- and post-hospital delay in managing stroke patients. Patients with acute stroke were enrolled in 2 tertiary hospitals in Jiangsu Province. On January 31, 2020, Chinese government announced several nation-wide strategies for preventing the COVID-19 pandemic. To evaluate the impacts of the pandemic on stroke management, patients with acute stroke within 2 months before and after this time point were analyzed in this study. Acute stroke was diagnosed based on clinical symptoms and CT or MRI scans. Patients who reached the hospitals within 7 days after stroke onset were included. All participants and their relatives provided written informed consent, and the study was approved by the ethics committees of the participated hospital.

### Treatment Delay Assessment

Demographic and clinical data were collected after hospitalization. Onset-to-door time (ODT) was defined as the duration from stroke symptom onset or time last known well to hospital arrival, which included awareness time, decision time and transporting time. Decision time is defined as the duration from symptom onset to the decision being made to go to hospital. Door-to-needle time (DNT) was defined as the time from hospital arrival to the start of intravenous thrombolysis. For those who did not meet the criteria of intravenous thrombolysis, DNT was based on a presumed thrombolytic therapy of earliest possibility. Door-to-puncture (DPT) time was defined as the duration from hospital arrival to groin puncture for mechanical thrombectomy. For those who did not meet the criteria of mechanical thrombectomy, DPT was based on a presumed mechanical thrombectomy of earliest possibility. Potential influencing factors for treatment delay, such as residency, means of transportation and level of the first visited hospital, were retrieved and analyzed. The severity of stroke was measured with National Institutes of Health Stroke Scale (NIHSS). In this study, we choose a 4.5 hours as the cut point for defining delayed treatment because that 4.5 hours is the accepted deadline for rt-PA intravenous thrombolysis at present [1].

### Statistical Analysis

Continuous variables were expressed as mean± standard deviation (SD) or median and interquartile range (IQR) as appropriate. Categorical variables were presented as frequency and percentage. Continuous variables with normal distribution were compared using the Student’s t-test. The Chi-square and Fisher’s exact tests were used for comparing categorical values. Multiple-variable stepwise logistic regression was used to determine the main influencing factors of treatment delay. A two sided *P* value of <0.05 was deemed as statistical significance. All statistical analyses were performed using SPSS 25.

## Results

A total of 267 patients were included during the described time frames, of which 161 (60.3%) were arrived before and 106 (39.7%) after the COVID-19 pandemic. The mean age of the included patients was 69.1± 11.3 years, and 167 (62.5%) of them were male. The median (IQR) NIHSS score at admission was 6 (3–13) in the Pre-COVID-19 group and 8 (5–16) in the Post-COVID-19 group (*P*=0.040). However, no significant differences concerning age, sex, residence, education level, stroke subtypes and comorbidities were detected between patients arrived before and after the COVID-19 pandemic (**Table 1**).

**Table 1.** Characteristics and treatment delay of stroke patients before and after COVID-19 pandemic

| Characteristics | Pre-COVID-19<br>(n=161) | Post-COVID-19<br>(n=106) | *P* Value |
| --- | --- | --- | --- |
| Age, year, mean | 69.5±11.1 | 70.1±12.2 | 0.334 |
| Male gender, n (%) | 97 (60.2) | 70(66.0) | 0.133 |
| Education, n (%) |  |  | 0.242 |
| Elementary education | 48 (29.8) | 35 (33.0) |  |
| Secondary education | 90 (55.9) | 59 (55.6) |  |
| Higher education | 23 (14.3) | 12 (11.4) |  |
| Residence, n (%) |  |  | 0.411 |
| Urban | 108 (67.1) | 75 (70.8) |  |
| Rural | 53 (32.9) | 31 (29.2) |  |
| Stroke subtype, n (%) |  |  | 0.532 |
| Ischemic stroke | 134 (83.2) | 91 (85.8) |  |
| Hemorrhagic stroke | 27 (16.8) | 15 (14.2) |  |
| NIHSS, median (IQR) | 6 (3-13) | 8 (5-16) | 0.040 |
| Stroke history, n (%) | 35 (21.7) | 24 (22.6) | 0.871 |
| Hypertension, n (%) | 103 (67.0) | 67 (63.2) | 0.642 |
| Diabetes, n (%) | 72 (44.7) | 50 (47.2) | 0.556 |
| Hyperlipidemia, n (%) | 53 (32.9) | 33 (31.1) | 0.734 |
| Atrial fibrillation, n (%) | 18 (11.1) | 14 (13.2) | 0.799 |
| Coronary heart disease, n (%) | 22 (13.6) | 18 (17.0) | 0.677 |
| Smoking, n (%) | 75 (46.6) | 43 (40.5) | 0.143 |
| Alcohol drinking, n (%) | 55 (34.1) | 40 (37.7) | 0.400 |
| ODT, median (IQR) min | 202 (25-492) | 317 (65-790) | 0.010 |
| Decision time | 129 (55-430) | 244 (80-710) | <0.001 |
| Transportation | 73 (31-93) | 67 (33-88) | 0.316 |
| DTN, median (IQR) min | 50 (40-75) | 65 (48-84) | 0.048 |
| Onset to treatment within 4.5h, % | 56 (34.8) | 31 (29.0) | 0.032 |
| Intravenous thrombolysis, %* | 27 (20.1) | 14 (15.4) | 0.030 |
| Mechanical thrombectomy, %* | 21 (15.7) | 11 (12.1) | 0.115 |

IQR indicates interquartile range; NIHSS, National Institutes of Health Stroke Scale;
ODT, onset-to door time; DTN, door-to-needle time.
\*Patients with ischemic stroke.

The ODT, a proxy of pre-hospital delay, was significantly longer in post-than pre-COVID-19 pandemic patients (317 [IQR 65-790] vs 202 [IQR 25-492] min; *P*=0.010).The decision time for patients after the COVID-19 pandemic was significantly longer than that of those before COVID-19 pandemic (129 [IQR 55-430] vs 244 [IQR 80-710] min, *P*<0.001). Time used for transportation was similar between patients before and after the COVID-19 pandemic (67 [IQR 33-88] vs 73 [IQR 31-93] min; *P*=0.316). DNT was prolonged significantly after the implementation of anti-pandemic strategies (65 [IQR 48-84] vs 50 [IQR 40-75] min, *P*=0.048). The proportion of patients from onset to treatment within 4.5 hours was significantly decreased after the COVID-19 pandemic (29.0% vs 34.8%, *P*=0.032). The proportion of patients with intravenous thrombolysis in those with acute ischemic stroke was decreased significantly after the pandemic (15.4% vs 20.1%; *P*=0.030). While the proportion of patients with mechanical thrombectomy in those with acute ischemic stroke remained unchanged (12.1% vs 15.7%, *P*=0.115, **Table 1**).

When compared with patients who arrived hospital within 4.5 hours (ONT ≤ 4.5 hours), those who arrived hospital latter (ONT >4.5h) had lower education level (elementary education: 18.4% vs 37.2%, *P*=0.018), more likely lived in rural areas (26.4% vs 33.9%, *P*=0.044), less likely had hemorrhagic stroke (18.4% vs 14.4%, *P*=0.041), had lower NIHSS scores (8 vs 3, *P*=0.031), less likely transferred by EMS (43.7% vs 16.1%, *P*<0.001), more likely had self-management after stroke onset (9.2% vs 87.8%, *P*<0.001), and more likely had stroke after the COVID-19 pandemic (27.6% vs 45.6%, *P*=0.031, **Table 2**).

**Table 2.** Influencing factors for delayed treatment

| Characteristics | Onset-to-needle time |  | *P* Value |
| --- | --- | --- | --- |
|  | ≤4.5 hour<br>(n=87) | >4.5 hour<br>(n=180) |  |
| Age, y, mean | 67.5±12.2 | 70.1±11.8 | 0.134 |
| Male gender, n (%) | 56 (64.4) | 111 (61.6) | 0.736 |
| Education, n (%) |  |  | 0.018 |
| Elementary education | 16 (18.4) | 67 (37.2) |  |
| Secondary education | 50 (57.5) | 99 (55.0) |  |
| Higher education | 21 (24.1) | 14 (7.8) |  |
| Residence, n (%) |  |  | 0.044 |
| Urban | 64 (73.6) | 119 (66.1) |  |
| Rural | 23 (26.4) | 61 (33.9) |  |
| Stroke subtype, n (%) |  |  | 0.041 |
| Ischemic stroke | 71 (81.6) | 154 (85.6) |  |
| Hemorrhagic stroke | 16 (18.4) | 26 (14.4) |  |
| NIHSS at admission, median (IQR) | 8 (3-14) | 3 (2-7) | 0.031 |
| Stroke history, n (%) | 21 (24.1) | 38 (21.1) | 0.523 |
| Hypertension, n (%) | 58 (66.6) | 112 (62.2) | 0.278 |
| Diabetes, n (%) | 39 (44.8) | 73 (40.6) | 0.298 |
| Hyperlipidemia, n (%) | 27 (31.0) | 59 (32.7) | 0.776 |
| Atrial fibrillation, n (%) | 13 (14.9) | 19 (10.6) | 0.165 |
| Coronary heart disease, n (%) | 14 (14.9) | 28 (15.6) | 0.679 |
| Current smoker, n (%) | 42 (48.3) | 76 (42.2) | 0.108 |
| Regular drinker, n (%) | 34 (39.1) | 61 (33.9) | 0.182 |
| Self-management after onset, n (%) | 8 (9.2) | 158 (87.8) | <0.001 |
| Transporting by EMS, n (%) | 38 (43.7) | 29 (16.1) | <0.001 |
| Post-COVID-19 period, n (%) | 24 (27.6) | 82 (45.6) | 0.035 |

IQR, interquartile range; NIHSS, National Institutes of Health Stroke Scale; EMS,
emergency medical services.

**Table 3** presents the potential influencing factors for delayed treatment (ONT>4.5h) by multivariate logistic regression analysis. Compared with patients before COVID-19 pandemic, patients after COVID-19 pandemic had an OR of 1.52 (95% CI, 1.02–2.94) for treatment delay. Compared with patients with advanced education, those with elementary education had an odds of 1.41 (95% CI, 1.08–2.31) for treatment delay. Compared with patients living in urban, those living in rural area had an odds of 1.20 (95% CI, 1.01–1.42) for treatment delay. Patients who firstly chose to self-manage stroke after onset had an OR of 2.03 (95% CI, 1.40–3.76) for treatment delay. Patients transported by EMS had an OR of 0.76 (95% CI, 0.68–0.86) for treatment delay. Patients with baseline NIHSS >10 had an OR of 0.64 (95% CI, 0.45–0.89) for treatment delay.

**Table 3.** Multivariate logistic regression analysis of influencing factors for delayed treatment (ONT>4.5h)

| Variables | OR | 95% CI | *P* |
| --- | --- | --- | --- |
| Elementary vs higher education | 1.41 | 1.08-2.31 | 0.045 |
| Rural vs urban residency | 1.20 | 1.01-1.42 | 0.030 |
| NIHSS > 10 vs ≤10 | 0.64 | 0.45-0.89 | <0.001 |
| Self-management after onset | 2.03 | 1.40-3.76 | <0.001 |
| Transporting by ambulance vs other means | 0.76 | 0.68-0.86 | 0.038 |
| Onset after COVID-19 pandemic | 1.52 | 1.02-2.94 | 0.010 |

ONT indicates onset-to-needle time; OR, odds ratio; CI, confidence interval; NIHSS, National Institutes of Health Stroke Scale.

## Discussion

The current study highlights the impact of COVID-19 pandemic on treatment delay in patients with acute stroke. ODT and DTN were significantly prolonged, and proportion of patients arrived within the time window for intravenous thrombolysis was significantly decreased after COVID pandemic.

During COVID-19, patients may be reluctant to seek medical help for fear of being infected. Patients with mild symptoms may stay at home and managing stroke be themselves or their relatives. This speculation was partly confirmed by the higher NIHSS score in post-COVID-19. A similar pattern of delay in seeking medical care due to fear of being infected within the hospitals was observed in the Ebola epidemic in West Africa [6].

During the COVID-19 pandemic, the onset to needle time was significantly prolonged than before. Traffic control during the pandemic may delay the patient transportation. Social distance may influence the management of stroke patients. Procedures for COVID-19 prevention, such as information inquiring concerning travel and contact history, temperature measuring, chest X-ray or CT scanning, coronavirus nucleic acid or antibody testing, blood cell counting, and multidisciplinary consultation may all delay the management of stroke. On the other hand, medical staff may need more time to wear protective devices before they could manage stroke patients during the COVID-19 pandemic.

This study associated higher NIHSS score with shorter pre-hospital delay. This is consistent with some previous studies [7, 8], but not with others [9]. Patients with severe symptoms may be more obvious to be identified, but severe symptoms may render patients from seeking for help when alone. Transferred with ambulance was associated with shorter pre-hospital delay [10-13]. Early awareness of stroke onset and rapid response are crucial to shorten the treatment delay [14, 15]. Previous studies [16] indicated that major factors for pre-hospital delay included unawareness of stroke symptoms, lack of understanding on importance of early response, and lack of knowledge on early management. Previous studies demonstrated that only 15.6% of respondents knew stroke warning signs [17]. A large proportion of respondents think that stroke symptoms may not warrant emergent management [18].

Several limitations should be addressed when interpreting the results of current study. Firstly, patients were enrolled outside the epicenter of COVID-19 pandemic in China, which may have under-estimated the impacts of the pandemic on stroke management. Secondly, with the accumulation of coping experiences, the impacts of COVID-19 pandemic on stroke management may be relieved. Finally, although the impacts of COVID-19 on intravenous thrombolysis and mechanical thrombectomy were analyzed, the impacts on stroke outcomes (e.g. 90-day modified Rankin Scale) were not analyzed.

In conclusion, COVID-19 pandemic has a remarkable influence on the management of acute ischemic stroke. Both pre- and post-hospital delays were prolonged significantly, and proportion of patient arrived within the 4.5-hour time window for intravenous thrombolysis treatment was decreased. Given that anti-COVID-19 measures are becoming medical routines, efforts are warranted to shorten the delay so that stroke outcome could be improved in this complex time.

### Statement of Ethics

Subjects (or their parents or guardians) have given their written informed consent for being treated for IV tPA. The article is exempt from ethical committee approval since IV tPA is considered the standard of care for treating AIS and there has been no disclosure of the patients’ information in this article.

## Data Availability

The data that support the findings of this study are available on request from the corresponding author.

## Funding Statement

The work was supported by [National Natural Science Foundation of China] grant numbers [81870947].

## Conflicts of Interest Statement

None declared.

## Author Contributions

SG and ZD: study design, interpretation of results and manuscript drafting. SG, YB and HS: study design and interpretation of results. SG, ZD, YB, HS and XZ: data collection. GX and SG: study design, statistical analysis and critical revision of manuscript. SG: interpretation of results and critical revision of manuscript. GX and XL have full access to all of the data in the study and take responsibility for the integrity of the data and the accuracy of the data analysis.

## Notes

### Competing Interest Statement

The authors have declared no competing interest.

### Funding Statement

This study was supported in part by the National Natural Science Foundation of China (NO.81870947).

### Author Declarations

This study was approved by the institutional review board of The Affiliated Yixing Hospital of Jiangsu University (IRB No.2020-001). All patients provided written informed consent.

